# Association of inflammatory markers with the severity of COVID-19

**DOI:** 10.1101/2020.04.14.20065680

**Authors:** Furong Zeng, Ying Guo, Mingzhu Yin, Xiang Chen, Guangtong Deng

## Abstract

**Background:** The ongoing worldwide epidemic of Coronavirus Disease 2019 (COVID-19), caused by severe acute respiratory syndrome coronavirus 2 (SARS-CoV- 2), has posed a huge threat to global public health. However, with regard to the effects of inflammatory markers on the severity of COVID-19, studies have reported associations that vary in strength and direction.

**Aims:** In the meta-analysis, we aimed to provide an overview of the association of inflammatory markers with severity of COVID-19.

**Methods:** The following databases were searched: PubMed, Embase, Cochrane Library, Wanfang database and CNKI (China National Knowledge Infrastructure) database until March 20, 2020. Weighted mean difference (WMD) and 95% confidence intervals (CIs) were pooled using random or fixed-effects models.

**Results:** A total of 16 studies were included in our analysis comprising of 3962 patients with COVID-19. Random-effects results demonstrated that patients with COVID-19 in non-severe group had lower levels for CRP (WMD = -41.78 mg/l, 95% CI = [-52.43, - 31.13], P < 0.001), PCT (WMD = -0.13 ng/ml, 95% CI = [-0.20, -0.05], P < 0.001), IL- 6 (WMD = -21.32 ng/l, 95% CI = [-28.34, -14.31], P < 0.001), ESR (WMD = - 8.40 mm/h, 95% CI = [-14.32, -2.48], P = 0.005), SAA (WMD = -43.35 μg/ml, 95% CI = [-80.85, -5.85], P = 0.020) and serum ferritin (WMD = -398.80 mg/l, 95% CI = [- 625.89, -171.71], P < 0.001), compared with those in severe group. Moreover, survivors had lower level for IL-6 than non-survivors with COVID-19 (WMD = -4.80 ng/ml, 95% CI = [-5.87, -3.73], P < 0.001). These results were consistent through sensitivity analysis and publication bias assessment.

**Conclusions:** The meta-analysis highlights the association of inflammatory markers with the severity of COVID-19. Measurement of inflammatory markers might help clinicians to monitor and evaluate the severity and prognosis of COVID-19.

## Introduction

The ongoing worldwide pandemic of Coronavirus Disease 2019 (COVID-19) has posed a huge threat to global public health (WHO, 2020a). The pathogen has been identified as a novel single-stranded ribonucleic acid (RNA) betacoronavirus named as severe acute respiratory syndrome coronavirus 2 (SARS-CoV-2), which share a great phylogenetic similarity with severe acute respiratory syndrome coronavirus (SARS-CoV)(Wu F. et al., 2020, Zhou P. et al., 2020). As of March, 21, 2020, a total of 266, 073 confirmed cases from 150 countries and territories were reported, including 11,183 deaths (WHO, 2020b). The COVID-19 represents a spectrum of clinical severity ranged from asymptomatic to critical pneumonia, acute respiratory distress syndrome (ARDS) and even death(Guan et al., 2020). Therefore, fully monitoring the severity of COVID- 19 and effective early intervention are the fundamental measures for reducing mortality. Accumulating evidence has suggested that inflammatory responses play a critical role in the progression of COVID-19, and several markers have some tracing and detecting accuracy for disease severity and fatality(Mehta et al., 2020, Stebbing et al., 2020, Wu C. et al., 2020). Inflammatory markers such as procalcitonin (PCT), serum ferritin, erythrocyte sedimentation rate (ESR), acute phase reactant protein C reactive protein (CRP) and interleukin-6 (IL-6) have been reported to be significantly associated with the high-risks of the development of severe COVID-19(Cheng et al., 2020, Gao et al., 2020, Qin et al., 2020). Moreover, increased levels of serum amyloid A(SAA) are shown to be involved in COVID-19 pathogenesis and may serve as a potential biomarker for monitoring disease progression(Cheng et al., 2020, Xiang et al., 2020). However, these results remain controversial due to no observed difference in the levels of IL-6, SAA, ESR and CRP by other studies (Chen L. et al., 2020, Wu C. et al., 2020, Zhang et al., 2020).

To our best of knowledge, the overall inflammatory profile is to-date missing due to the insufficient sample size. Here we performed an analysis of the current scientific literature to provide an overview of the association of inflammatory markers with severity in patients with COVID-19.

## Methods

### Search strategy

This meta-analysis was performed according to the Preferred Reporting Items for Systematic Reviews and Meta-Analyses (PRISMA) guidelines. Original studies reporting COVID-19 were searched until March 20, 2020 through PubMed, Embase, Cochrane Library, Wanfang database and CNKI (China National Knowledge Infrastructure) database. The following combined search terms were used: (“novel coronavirus” OR “Wuhan virus” OR “Chinese virus” OR “nCoV-2019” OR “2019- nCoV” OR “COVID-19” OR “SARS-CoV-2”). All eligible articles were retrieved, and their references of identified publications were searched for further potentially relevant articles(Gao et al., 2020, Zhou F. et al., 2020).

### Selection criteria

English-language or Chinese-language publications reporting concentrations of inflammation markers in patients with COVID-19 were included if they met the following criteria: (1) patients could be grouped into severe COVID-19 and non-severe COVID-19, or non-intensive care unit (ICU) group and ICU group, or survivors and non-survivors with COVID-19; (2) literature sources and necessary data were available; and (3) the diagnostic criteria of COVID-19 were clarified based on the laboratory-confirmed SARS-CoV-2 infection. If there were two or more studies from the same authors or institutions, only the study with the largest sample size was chosen. Studies were excluded if full text of the trial was not available or they did not fulfill the inclusion criteria.

### Data extraction and quality assessment

The records from the initial search were scanned by two authors to exclude any duplicate and irrelevant studies. The following data were extracted: first authors, publication date, country of origin, grouping situation, cases, age, sex and levels of inflammatory markers in different groups. Stratified data or interquartile range (IQR) were converted to mean (±SD) using mathematical formulas for meta-analysis(Luo et al., 2018, Wan et al., 2014). Any discrepancies were resolved by discussion. Quality assessments of all potentially eligible studies were conducted using the Newcastle- Ottawa Scale (NOS). There are eight items in three aspects: selection, comparability and exposure. The full score was 10 stars. Four to six stars was regarded as a moderate-quality study, and seven to nine stars as a high-quality study. Studies with NOS scores lower than 7 were recognized to be of inferior quality and therefore excluded.

### Statistical analysis

All the statistical analyses were carried out by STATA (Version 12.0; STATA Corporation, College Station, TX, USA) software. Statistical heterogeneity was assessed with I^2^ and P-value. A fixed effects model was adopted without significant heterogeneity (I^2^ ≤ 50% and P ≥ 0.1), while a random effects model was employed in all other instances (I^2^ > 50% or P < 0.1)(Zeng et al., 2019, Zeng et al., 2020). Weighted mean difference (WMD) with 95% confidence intervals (95% CI) was calculated for inflammatory markers. Sensitivity analysis was performed by omitting one study each time through influence analysis to assess the stability of results. Besides, standard mean difference (SMD) were used to explore the consistence of the conclusion. Publication bias was evaluated by Egger’s test. If publication bias was conformed, the Duval’s trim and fill method was implemented to adjust for this bias. P < 0.05 was considered statistically significant.

## Results

### Literature search and studies characteristics

The initial literature search generated altogether 5384 records with 734 studies subsequently excluded due to duplication (**Figure 1**). After a review of the titles and abstracts, we obtained 35 studies by excluding an additional 4615 studies. We further excluded 19 studies by scanning the full text which did not report inflammatory markers. Finally, 16 studies were included in our analysis(Chang et al., 2020, Chen C. et al., 2020, Chen L. et al., 2020, Cheng et al., 2020, Fang et al., 2020, Gao et al., 2020, Huang et al., 2020, Li et al., 2020, Peng et al., 2020, Qin et al., 2020, Ruan et al., 2020, Wu C. et al., 2020, Xiang et al., 2020, Xiao et al., 2020, Zhang et al., 2020, Zhou F. et al., 2020). Characteristics of 16 eligible studies were presented in **Table 1**. All these studies come from China and were published in 2020, involving 3962 patients. 12 studies were grouped by non-severe and severe groups, 2 studies grouped by non-ICU and ICU groups, and 2 studies grouped by survivors and non-survivors with COVID-19. Obviously, patients in severe group, ICU group or non-survivors group had older age than those in corresponding control group. There were no obvious differences in the sex distribution of patients for each study. All studies were deemed of high quality with 7 or more NOS scores and details could be found in **Table 2**.

**Table 1.** Characteristics of enrolled studies.

| First author | Year | Country | Groups | Cases | Age | Sex (male, %) | Inflammatory markers | Quality |
| --- | --- | --- | --- | --- | --- | --- | --- | --- |
| Chang Z | 2020 | China | Nonsevere | 93 | - | - | CRP, PCT, IL-6 | 8 |
|  |  |  | Severe | 57 |  |  |  |  |
| Chen C | 2020 | China | Nonsevere | 126 | 57.0±15.6 | 66 (52.3) | CRP | 7 |
|  |  |  | Severe | 24 | 68.5±13.6 | 18 (75) |  |  |
| Chen L | 2020 | China | Nonsevere | 15 | - | - | IL-6, CRP | 7 |
|  |  |  | Severe | 14 |  |  |  |  |
| Cheng K | 2020 | China | Nonsevere | 282 | 49.7±11.9 | 145(51.42) | IL-6, CRP, PCT, ESR, SAA, Serum ferritin | 8 |
|  |  |  | Severe | 181 | 54.7±13.5 | 99(54.7) |  |  |
| Fang X | 2020 | China | Nonsevere | 55 | 39.9±14.9 | 27 (49.1) | CRP | 7 |
|  |  |  | Severe | 24 | 56.7±14.4 | 18 (75) |  |  |
| Gao Y | 2020 | China | Nonsevere | 28 | 43.0±14.0 | 17 (60.7) | IL-6, CRP, PCT, fibrinogen | 9 |
|  |  |  | Severe | 15 | 45.2±7.7 | 9 (60.0) |  |  |
| Jin Z | 2020 | China | Nonsevere | 82 | 51.6±10.7 | 38 (46.3) | CRP, PCT, SAA | 7 |
|  |  |  | Severe | 56 | 62.7±13.6 | 33 (56.9) |  |  |
| Li K | 2020 | China | Nonsevere | 58 | 41.9±10.6 | 29 (50) | CRP, PCT | 7 |
|  |  |  | Severe | 25 | 53.7±12.3 | 15 (60) |  |  |
| Peng Y | 2020 | China | Nonsevere | 96 | 61.5±9.4 | 44 (45.83) | CRP, PCT | 8 |
|  |  |  | Severe | 16 | 58.2±7.3 | 9 (56.25) |  |  |
| Qin C | 2020 | China | Nonsevere | 166 | 52.0±15.5 | 80 (48.2) | IL-6, CRP, PCT, ESR, Serum ferritin | 7 |
|  |  |  | Severe | 286 | 60.3±13.4 | 155 (54.2) |  |  |
| Wu C | 2020 | China | Nonsevere | 117 | 47.3±10.5 | 68 (58.1) | IL-6, CRP, ESR, Serum ferritin | 7 |
|  |  |  | Severe | 84 | 59.2±14.3 | 60 (71.4) |  |  |
| Xiang T | 2020 | China | Nonsevere | 40 | 40.6±14.3 | 25 (63.5) | CRP, PCT, ESR, SAA | 8 |
|  |  |  | Severe | 9 | 53.0±14.0 | 8 (88.9) |  |  |
| Xiao K | 2020 | China | Nonsevere | 107 | 43.05±1.13 | 52 (48.6) | IL-6, CRP, PCT | 7 |
|  |  |  | Severe | 36 | 51.28±5.58 | 20 (55.6) |  |  |
| Huang C | 2020 | China | Non-ICU | 28 | 49.2±12.9 | 19 (68) | PCT | 9 |
|  |  |  | ICU | 13 | 50.5±16.6 | 11 (85%) |  |  |
| Ruan Q | 2020 | China | Survivor | 82 | 51.6±7.6 | 53 (65) | IL-6, CRP, | 7 |
|  |  |  | Non-survivor | 68 | 64.3±14.0 | 49 (72) |  |  |
| Zhou F | 2020 | China | Survivor | 137 | 51.6±9.7 | 81 (59) | IL-6, PCT, Serum ferritin | 7 |
|  |  |  | Non-survivor | 54 | 69.4±9.9 | 38 (70) |  |  |

CRP: acute phase reactant protein C reactive protein; PCT: procalcitonin; IL-6: interleukin-6; ESR: erythrocyte sedimentation rate; SAA: serum amyloid A.

**Table 2.** Methodological quality of enrolled studies based on Newcastle-Ottawa Scale (NOS).

| Included studies | Year | Is the definition adequate? | Representativeness of the cases | Selection of controls | Definition of controls | Comparability of both groups | Ascertainment of diagnosis | Same ascertainment method for both groups | Nonresponse rate | Total scores |
| --- | --- | --- | --- | --- | --- | --- | --- | --- | --- | --- |
| Chang Z | 2020 | ☆ | ☆ | ☆ | ☆ | ☆ | ☆ | ☆ | ☆ | 8 |
| Chen C | 2020 | ☆ | ☆ | ☆ | ☆ | - | ☆ | ☆ | ☆ | 7 |
| Chen L | 2020 | ☆ | ☆ | ☆ | ☆ | - | ☆ | ☆ | ☆ | 7 |
| Cheng K | 2020 | ☆ | ☆ | ☆ | ☆ | ☆ | ☆ | ☆ | ☆ | 8 |
| Fang X | 2020 | ☆ | ☆ | ☆ | ☆ | - | ☆ | ☆ | ☆ | 7 |
| Gao Y | 2020 | ☆ | ☆ | ☆ | ☆ | ☆ ☆ | ☆ | ☆ | ☆ | 9 |
| Jin Z | 2020 | ☆ | ☆ | ☆ | ☆ | - | ☆ | ☆ | ☆ | 7 |
| Li K | 2020 | ☆ | ☆ | ☆ | ☆ | - | ☆ | ☆ | ☆ | 7 |
| Peng Y | 2020 | ☆ | ☆ | ☆ | ☆ | ☆ | ☆ | ☆ | ☆ | 8 |
| Qin C | 2020 | ☆ | ☆ | ☆ | ☆ | - | ☆ | ☆ | ☆ | 7 |
| Wu C | 2020 | ☆ | ☆ | ☆ | ☆ | - | ☆ | ☆ | ☆ | 7 |
| Xiang T | 2020 | ☆ | ☆ | ☆ | ☆ | ☆ | ☆ | ☆ | ☆ | 8 |
| Xiao K | 2020 | ☆ | ☆ | ☆ | ☆ | - | ☆ | ☆ | ☆ | 7 |
| Huang C | 2020 | ☆ | ☆ | ☆ | ☆ | ☆ ☆ | ☆ | ☆ | ☆ | 9 |
| Ruan Q | 2020 | ☆ | ☆ | ☆ | ☆ | - | ☆ | ☆ | ☆ | 7 |
| Zhou F | 2020 | ☆ | ☆ | ☆ | ☆ | - | ☆ | ☆ | ☆ | 7 |

**Figure 1.**
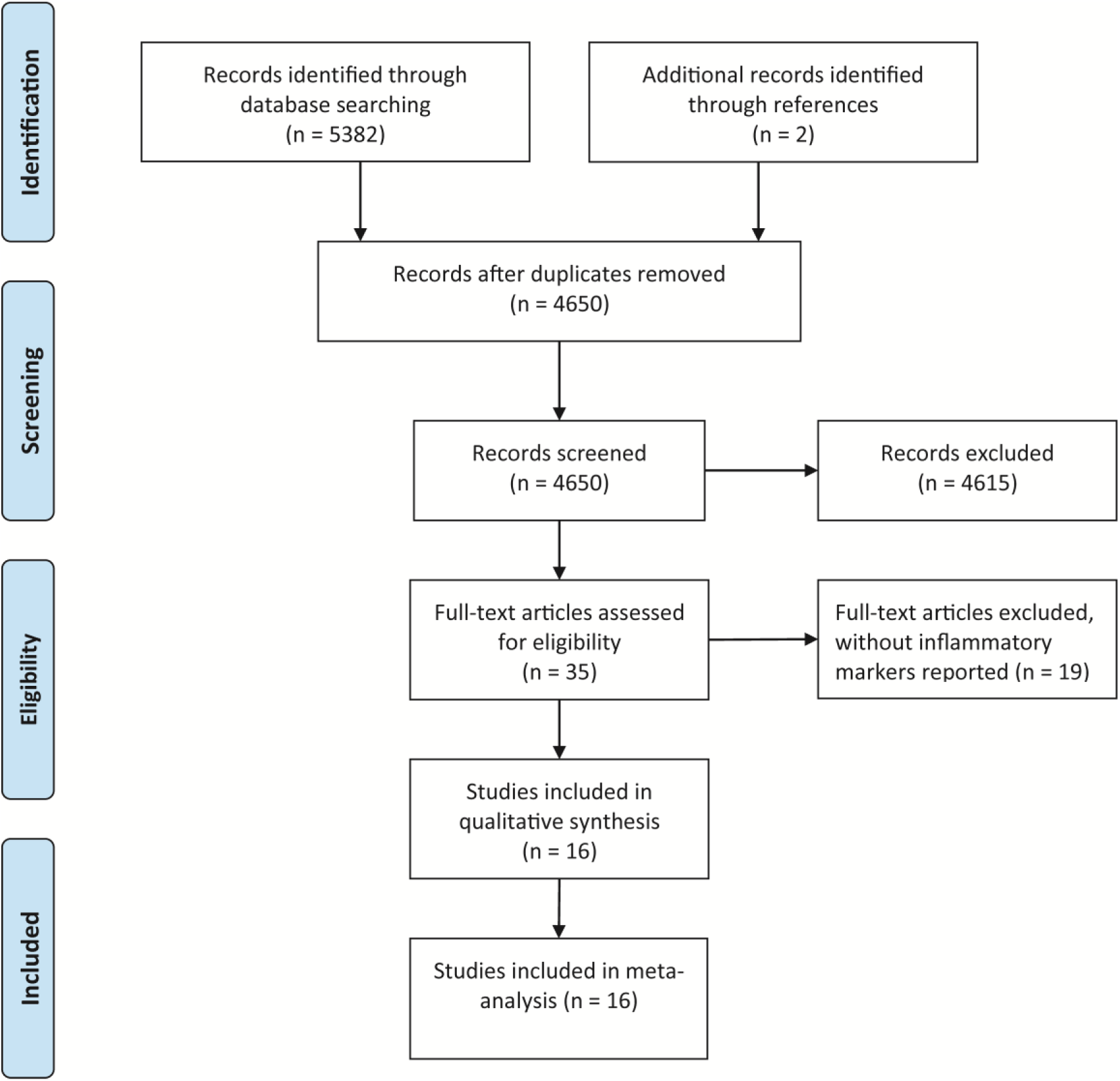
Literature search and filtering of studies.

### Association of inflammatory markers with the severity of COVID-19

For the patients stratified by severity of COVID-19, random-effects results demonstrated that compared with patients in severe group, patients in non-severe group had lower levels for CRP (WMD = -41.78 mg/l, 95% CI = [-52.43, -31.13], P < 0.001), PCT (WMD = -0.13 ng/ml, 95% CI = [-0.20, -0.05], P < 0.001), IL-6 (WMD = - 21.32 ng/l, 95% CI = [-28.34, -14.31], P < 0.001), ESR (WMD = -8.40 mm/h, 95% CI = [-14.32, -2.48], P = 0.005), SAA (WMD = -43.35 μg/ml, 95% CI = [-80.85, -5.85], P = 0.020) and serum ferritin (WMD = -398.80 mg/l, 95% CI = [-625.89, -171.71], P < 0.001) (**Figure 2A-2E**). Besides, there are two studies grouped by survivors and non-survivors with COVID-19 reporting the level of IL-6, and fixed-effect result arrived at a similar conclusion that survivors had lower level for IL-6 than non-survivors with COVID-19 (WMD = -4.80 ng/ml, 95% CI = [-5.87, -3.73], P < 0.001) (**Figure 2F**).

**Figure 2.**
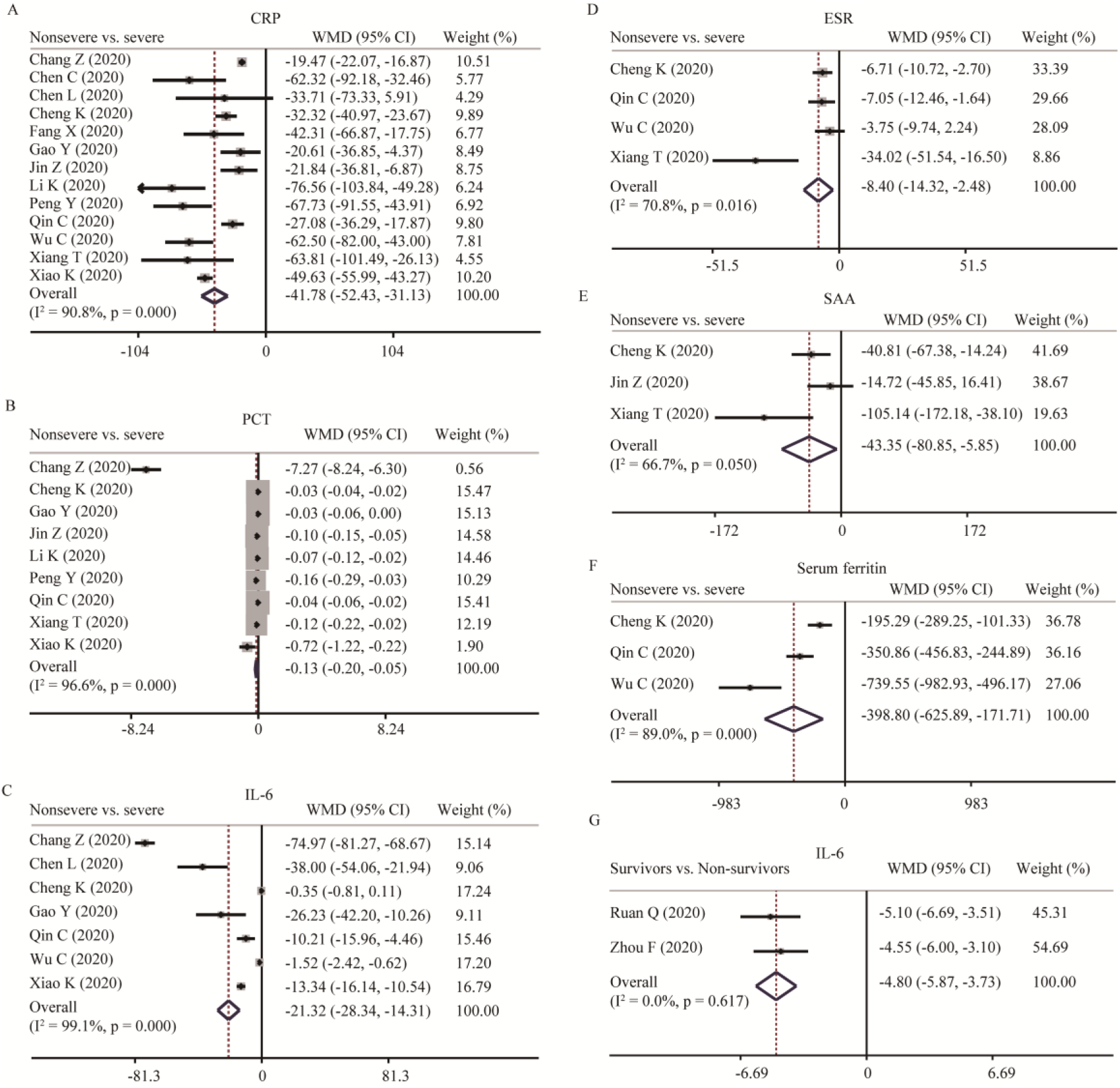
Forest plot in different subgroups. A-F. Forest plot between non-severe and severe groups for levels of CRP (A), PCT (B), IL-6 (C), ESR (D), SAA (E), and serum ferritin (F); G. Forest plot between survivors group and non-survivors group for levels of IL-6.

Additionally, one study on the level of fibrinogen between non-severe group and severe group, one study on the level of PCT between non-ICU group and ICU group, one study on the level of CRP and PCT between non-survivors and survivors, were not included in the meta-analysis due to their inadequate data; however, the results reported by these studies were consistent with the pooled results of our meta-analyses.

### Investigation of heterogeneity

Strong evidence of heterogeneity was found in all the comparisons (**Figure 2**). Sensitivity analyses indicated that the results were not influenced by excluding any one specific study in CRP, PCT, IL-6 and ESR between non-severe and severe groups (**Figure 3A-3D**). As for SAA and serum ferritin, the conclusions changed when deleting Cheng K’ study and Qin C’s study, separately, while the heterogeneity become larger, suggesting that it is better to keep these studies in the meta-analysis (**Figure 3E and 3F**). Egger’s test was conducted to evaluate the publication bias (**Table 3**). No significant publication bias was detected in most of the studies except for CRP (P = 0.012) and PCT (P = -0.036). When applying the trim-and-fill method, there were not any trials trimmed and filled in CRP. About PCT, after filling one trial, the revised result was still consistent using random model (WMD = − 0.169 ng/ml, 95% CI = [-0.255, - 0.084], P < 0.001) or fixed model (WMD = − 0.062 ng/ml, 95% CI = [− 0.072, − 0.053], P < 0.001). Besides, using standard mean difference (SMD) for the meta-analysis still did not change the conclusions (**Table 4**).

**Table 3.** publication bias assessment.

| Outcome | Studies | P value (Egger's test) |
| --- | --- | --- |
| CRP | 13 | 0.016 |
| PCT | 9 | 0.036 |
| IL-6 | 7 | 0.012 |
| ESR | 4 | 0.185 |
| SAA | 3 | 0.468 |
| Serum ferritin | 3 | 0.243 |

CRP: acute phase reactant protein C reactive protein; PCT: procalcitonin; IL-6: interleukin-6; ESR: erythrocyte sedimentation rate; SAA: serum amyloid A.

**Table 4.** The results of the meta-analysis based on standard mean difference (SMD).

| Outcome | Studies | Participants | Heterogeneity |  | Model | SMD | 95% CI | P |
| --- | --- | --- | --- | --- | --- | --- | --- | --- |
|  |  |  | I<sup>2</sup> | P |  |  |  |  |
| Nonsevere vs. Severe |  |  |  |  |  |  |  |  |
| 1. CRP | 13 | 2092 | 95% | <0.001 | Random | -1.48 | [-1.95, -1.00] | <0.001 |
| 2. PCT | 9 | 1633 | 94% | <0.001 | Random | -1.11 | [-1.58, -0.63] | <0.001 |
| 3. IL-6 | 7 | 1481 | 98% | <0.001 | Random | -1.54 | [-2.38, -0.71] | <0.001 |
| 4. ESR | 4 | 1165 | 67% | 0.03 | Random | -0.34 | [-0.57, -0.10] | 0.005 |
| 5. SAA | 3 | 650 | 73% | 0.03 | Random | -0.41 | [-0.82, -0.00] | 0.050 |
| 6. Serum ferritin | 3 | 1116 | 80% | 0.007 | Random | -0.62 | [-0.90, -0.34] | <0.001 |
| Nonsurvivors vs. Survivors |  |  |  |  |  |  |  |  |
| IL-6 | 2 | 341 | 9% | 0.30 | Fixed | -1.23 | [-1.47, -0.98] | <0.001 |

CRP: acute phase reactant protein C reactive protein; PCT: procalcitonin; IL-6: interleukin-6; ESR: erythrocyte sedimentation rate; SAA: serum amyloid A.

**Figure 3.**
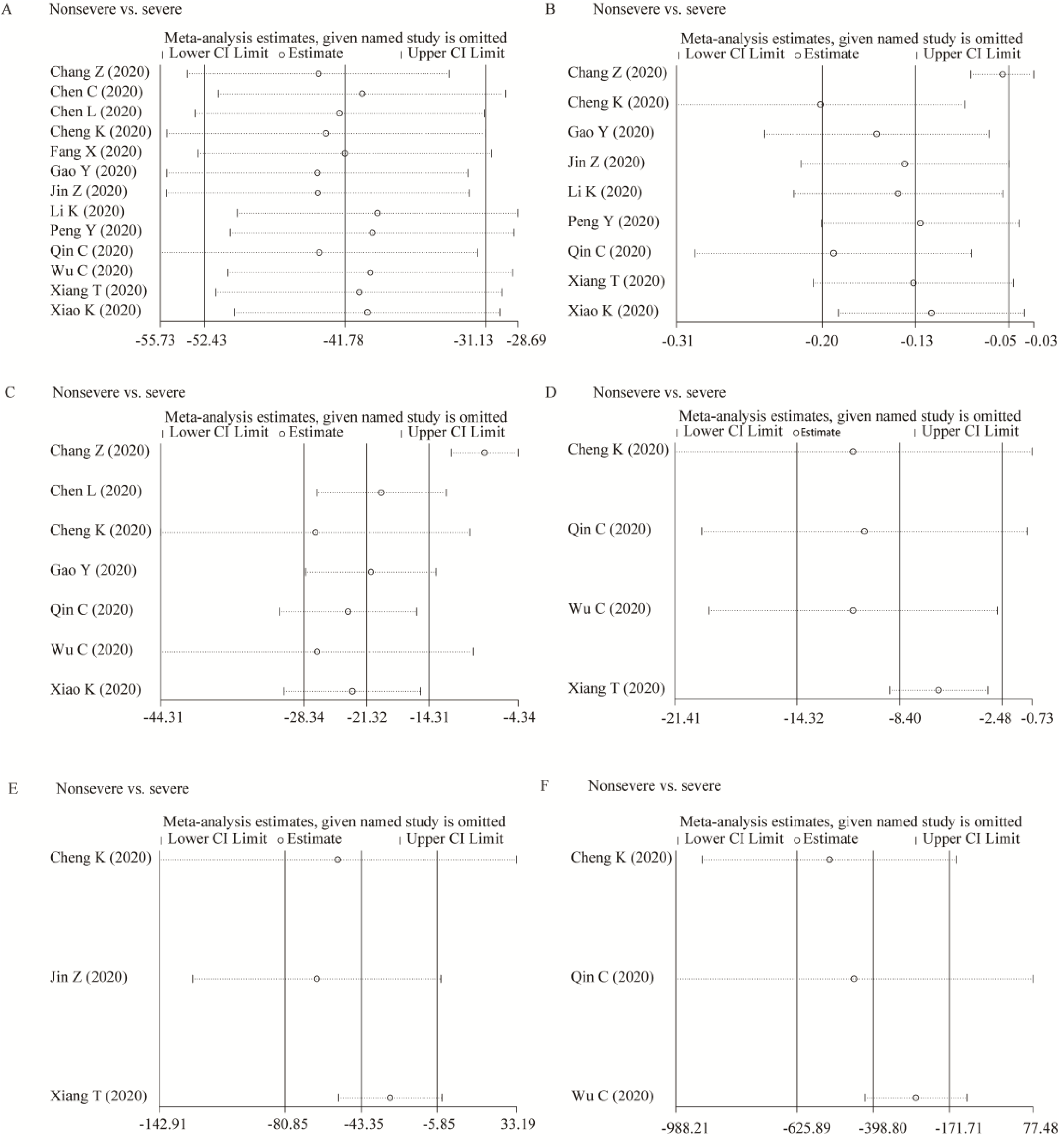
Sensitivity analyses. A-F. Sensitivity analyses between non-severe and severe groups for levels of CRP (A), PCT (B), IL-6 (C), ESR (D), SAA (E), and serum ferritin (F).

## Discussion

COVID-19), caused by SARS-CoV-2, is rapidly expanding worldwide. Despite most of cases have mild symptoms and good prognosis, COVID-19 can develop to ARDs and even death. To date, there is no effective therapy for COVID-19(Li and De Clercq, 2020, Russell et al., 2020). Therefore, it’s imperative to identify the markers monitoring the progression of disease and treat patients early.

Several studies have shown increased proinflammatory cytokines in serum of COVID- 19 patients. Also, anti-inflammatory agents for COVID-19 therapy highlights the critical role of inflammation in the progression of COVID-19(Mehta et al., 2020, Stebbing et al., 2020). However, the role of inflammatory markers in monitoring the severity of COVID-19 is still controversial. In this study, through analyzing the 16 retrospective studies, we concluded that inflammatory markers especially CRP, PCT, IL-6 and ESR were positively correlated with the severity of COVID-19.

IL-6 has been implicated in the 2003 SARS outbreak and the H5N1 avian influenza infections(Law et al., 2005, Saito et al., 2018). Recent studies showed that IL-6 and granulocyte-macrophage colony stimulating factor (GM-CSF) could be secreted by the active pathogenic T cell upon SARS-CoV-2 infection. And also, CD14+CD16+ inflammatory monocyte activated by GM-CSF could secret more IL-6 and other inflammatory factors(Zhou Yonggang et al., 2020). According to the New Coronavirus Pneumonia Prevention and Control Program (7th edition) published by the National Health Commission of China, decreasing level of IL-6 indicates the deterioration of COVID-19. Our study firstly provided an evidence-based medicine evidence through meta-analysis. Moreover, the level of IL-6 could not be routinely detected in many hospitals of China, but some inflammatory markers such as CRP, PCT and ESR usually could be detected. Our study firstly put forward that besides IL-6, other inflammatory markers such as CRP, PCT and ESR were also positively correlated with the severity of COVID-19. These conclusions were consistent through sensitivity analysis and publication bias assessment.

CRP is an exquisitely sensitive systemic marker of acute-phase response in inflammation, infection, and tissue damage, which could be used as indicator of inflammation(Pepys and Hirschfield, 2003). In Chen L et al. study, although no statistically significant difference was found in the level of CRP between non-severe and severe group, the mean level of CRP was higher in severe group than non-severe group(Chen L. et al., 2020). Other studies all reported CRP level was positively related to the severity of COVID-19. PCT is also a main inflammatory marker routinely measured in clinical practice. Among 9 studies, the levels of PCT were all higher in severe group than non-severe group. ESR is a non-specific inflammatory marker, which mainly reflects the changes of plasma protein types(Wu et al., 2018). In our meta-analysis, we found higher ESR level in severe group than non-severe group. One reason is that patients in severe group had higher inflammation. Another possible explanation is that patients with older age in severe group contributed to the higher level of ESR considering that the level of ESR increased with age(Piva et al., 2001).

We also found patients with COVID-19 in severe group had higher levels of SAA and serum ferritin than those in non-severe group. Considering that only three studies reported their levels and sensitivity analysis changed the conclusion, we temporarily could not conclude their association with the severity of COVID-19. SAA is a sensitive acute response protein which was used as a sensitive index to reflect the control of infection and inflammation. Serum ferritin is a surrogate marker of stored iron and increases in inflammation, liver disease, and malignancy(Cohen et al., 2010, Facciorusso et al., 2014, Kowdley et al., 2012). All of these highlights that overexuberant inflammatory response is associated with severity of COVID-19.

To our knowledge, this is the first meta-analysis on the associations of a series of inflammatory markers with the severity of COVID-19. Admittedly, our meta-analysis had some limitations. Firstly, noticeable heterogeneity exists in most of the analyses. Sensitivity analysis and SMD were used for the meta-analysis, yet the heterogeneity could not be eliminated completely. Secondly, reporting and publication bias may result from the lack of information or unpublished negative studies though the conclusion did not change through the trim-and-fill method. Thirdly, the studies included in our meta-analysis were mainly from China and whether the conclusion was consistent in other countries needs to be further investigated. Finally, it is underpowered to investigate the underlying mechanism of these inflammatory markers with the severity of COVID-19. In conclusion, inflammatory markers especially CRP, PCT, IL-6 and ESR were positively correlated with the severity of COVID-19. The association of SAA and serum ferritin with the severity of COVID-19 needs further clarified. Measurement of inflammatory markers might help clinicians to monitor and evaluate the severity and prognosis of COVID-19.

## Data Availability

All the data are available from the corresponding author on request.

## Authors contribution

FZ and GD were responsible for study design and writing; YG collected the data; GD analyzed the data; MY and XC revised the manuscript.

## Ethics in publishing

Approval was not required.

